# Clinical validation and performance evaluation of the automated Vitros Total Anti-SARS-CoV-2 Antibodies assay for screening of serostatus in COVID-19

**DOI:** 10.1101/2020.06.09.20126474

**Authors:** Emily Garnett, Joanna Jung, Estella Tam, Deepthi Rajapakshe, Stephen Cheney, Cameron Brown, Kenneth L. Muldrew, Jing Cao, Ila Singh, James Versalovic, Sridevi Devaraj

**Author notes:** Corresponding Author: Sridevi Devaraj, Ph.D., DABCC, FRSC, Professor of Pathology and Immunology, Baylor College of Medicine, Director, Clinical Chemistry, Texas Children’s Hospital, 6621 Fannin St. Houston, TX 77030.

## Abstract

**Objectives:** Evaluation of serostatus against SARS-CoV-2 has emerged as an important tool in identification of exposure to COVID-19. We report on the validation of the Vitros Anti-SARS-CoV-2 Total (CoV2T) assay for qualitative serological testing of SARS-CoV-2 antibodies.

**Methods:** We performed validation studies according to COLA guidelines, using samples previously tested for SARS-CoV-2 by RT-PCR. We evaluated precision, analytical interferences, and cross-reactivity with other viral infections. We also evaluated concordance with molecular and other serological testing, and evaluated seroconversion.

**Results:** The Vitros CoV2T assay exhibited acceptable precision, was resistant to analytical interference, and did not exhibit cross-reactivity with samples positive for other respiratory viruses. The CoV2T assay exhibited 100% negative predictive agreement (56/56) and 71% positive predictive agreement (56/79) with RT-PCR across all patient samples, and was concordant with other serological assays. Concordance with RT-PCR was 97% > 7 days after symptom onset.

**Conclusions:** The Vitros CoV2T assay was successfully validated in our laboratory. We anticipate it will be a useful tool in screening for exposure to SARS-CoV-2, however, the use of the CoV2T and other serological assays in clinical management of COVID-19 patients is yet unknown, and must be evaluated in future studies.

**Key points:** *What issue or core problem does the study address?:*

- Multiple serological assays for detection of anti-SARS-CoV-2 antibodies have received FDA Emergency Use Authorizations, but few data have been published on the performance of these assays.

*What would one take-home point for the working medical professional be?:*

- The Vitros Anti-SARS-CoV-2 Total assay is a total antibody test to be used as a serological screen for exposure to COVID-19. This assay performs well, and is comparable to other serological tests.

*What is the most significant or most interesting finding of the study?:*

- We confirmed that the Vitros Anti-SARS-CoV-2 Total assay, like other serological tests, is not suitable for diagnosis of acute infection, as it is not sensitive to infection <7 days post-onset.

## Introduction

SARS-CoV-2 is a novel coronavirus first identified in China in late 2019. The resultant disease, COVID-19, yields a range of presentations, from mild disease to severe pneumonia requiring mechanical ventilatory support, as well as thrombotic strokes, multi-inflammatory syndrome, and others ^1^. COVID-19 has since become a global pandemic and is continuing to spread, producing substantial morbidity, mortality, and economic impact ^2^. Accurate epidemiological information is important for the development of effective public health measures for containment and mitigation of COVID-19, but to date, case identification has been complicated by the wide range of clinical presentations of COVID-19. The mainstay of diagnostic testing is RT-PCR to identify SARS-CoV-2 viral RNA in respiratory swab specimens, but preanalytical (such as nasopharyngeal swab sampling) as well as analytical challenges have limited the utility of molecular testing as a means to screen for exposure to SARS-CoV-2 ^3,4^. The abundance of detectable nucleic acid also decreases over time, and other studies have suggested the RNA positive rate may decline to under 30% by 3 weeks after symptom onset ^5^. Consequently, the extent of COVID-19 infection remains unknown in many populations, limiting the ability to assess case fatality rates and hampering efforts to effectively quarantine infected individuals and limit spread.

Serological testing is used in the identification and management of many infectious diseases to provide evidence that an individual has had exposure to a pathogen and mounted an immune response. The laboratory workflow for serological testing is typically simpler and faster as well as less expensive than that of molecular testing, and specimen collection is more reproducible, which offers advantages for widespread screening. Serological testing has been suggested as a means of surveillance to determine actual numbers of COVID-19 infections, which can subsequently inform public health strategies. Previous studies have indicated that seropositivity in COVID-19 begins to occur approximately 7 days after symptom onset, although how long seropositivity remains after recovery and to what extent it indicates immunity to reinfection is not yet established ^6^.

Recommendations from the CDC and FDA indicate the use of total (IgG and IgM) anti-SARS-CoV-2 antibodies as evidence of previous viral exposure ^7^. Only a few serological assays have been granted Emergency Use Authorization (EUA) by the FDA for this purpose, but limited data are available on the analytical or clinical performance of these tests. Here, we describe validation of one of the first assays to receive EUA on an automated platform, the Vitros Anti-SARS-CoV-2 Total antibody assay, for screening of previous exposure to SARS-CoV-2 in our patient population.

## Materials and Methods

The Vitros (VITROS®) Anti-SARS-Cov-2 Total assay (CoV2T, Ortho Clinical Diagnostics, Raritan, NJ) detects total IgG and IgM directed against SARS-Cov-2, and was evaluated for use on the Vitros 5600 automated chemistry analyzer (Ortho Clinical Diagnostics, Raritan, NJ). The CoV2T assay uses a solid-phase SARS-CoV-2 spike protein antigen to capture antibodies in the patient specimen, and horseradish peroxidase-labeled recombinant SARS-CoV-2 antigen as a detection reagent. The assay is qualitative, and reports results as reactive or non-reactive, using a manufacturer-defined signal/cutoff (s/c) ratio of 1.00 as the decision limit. Numerical values for s/c are provided by the instrument, but not used in patient reports.

Specimens for validation were obtained under informed consent from healthy volunteers and from known COVID-19 patients, under an approved protocol from the Baylor College of Medicine Institutional Review Board (H47459). Known positive patients were previously diagnosed with COVID-19 by RT-PCR methods at Texas Children’s Hospital clinical laboratories, or at other institutions in the Texas Medical Center. Patient specimens were collected by venipuncture into K_2_EDTA tubes or serum separator tubes and processed upon receipt by the laboratory, with plasma or serum stored at 4 °C until analysis.

Precision studies were performed using a vendor-provided positive and negative control as well as a known negative sample and a known positive patient sample. Intra-assay precision was assessed by running the manufacturer-provided positive and negative control as well as one positive and negative sample 10 times in a single run, and inter-assay precision was assessed by running these samples 10 times on separate runs. Precision was assessed as % coefficient of variation (CV), using numerical s/c values provided by the instrument.

Accuracy studies were performed using 57 healthy volunteers with no known exposure, travel history, or symptoms of COVID-19 and 79 patient samples that were RT-PCR positive for SARS CoV-2 and/or admitted to the COVID ICU. Samples were tested on different days, by different operators. Accuracy was assessed as concordance with the positive or negative status of the specimen as assessed by molecular (RT-PCR) testing. These specimens were also tested for concordance with a semiquantitative SARS-CoV-2 IgG and IgM assay (SARS-CoV2 IgG or IgM ELISA, Ansh Laboratories, Webster, TX) on the Dynex DS2 (Dynex Technologies, Chantilly, VA), using the manufacturer’s cutoff value as a threshold for positive or negative results. An additional twelve samples were split and tested for concordance with results from a reference laboratory (Viracor Eurofins IgG and IgM panel).

Seroconversion in our patient population was assessed by correlation of chart review of 55 patients known to be positive for SARS-CoV-2 by RT-PCR and known date of symptom onset with sample reactivity by the CoV2T assay. Specimens were grouped by the number of days elapsed since the first reported symptom per patient history. Sensitivity was determined in samples that were post day 7 of symptom onset.

Analytical specificity was assessed by testing 14 different patient samples known to be positive for other viruses by molecular testing (including Influenza A, Influenza B, respiratory syncytial virus (RSV), adenovirus, rhinovirus, or other coronaviruses), but negative for SARS-CoV-2 by RT-PCR.

Interference testing was also performed by spiking known negative or positive samples with known concentrations of hemoglobin, conjugated bilirubin, and triglyceride-rich lipid (Sun Diagnostics, New Gloucester, ME). Results were considered acceptable if the spiked sample reactivity was concordant with the neat sample.

The effect of tube type was assessed by collecting specimens from five volunteers into serum separator or K_2_EDTA tubes, and measuring the resultant serum or plasma on the CoV2T assay. Concordance between tube types was assessed, with results considered acceptable if concordant.

Statistical analysis was performed in EP Evaluator. Results are given as mean ± standard deviation.

## Results

The Vitros CoV2T assay exhibited good precision. The CoV2T is a qualitative assay and does not report numerical results for patient use, but we examined the coefficient of variation using the signal to cutoff ratios (s/c). In intra-assay experiments, a known negative specimen was reported as nonreactive over 10 replicates, with s/c CV of 10.9%. A known positive specimen was reported as reactive, with s/c CV of 1.6%. Inter-assay experiments yielded CV of 9.7% for a negative specimen and 3.3% for a positive specimen (Table 1).

**Table 1.**
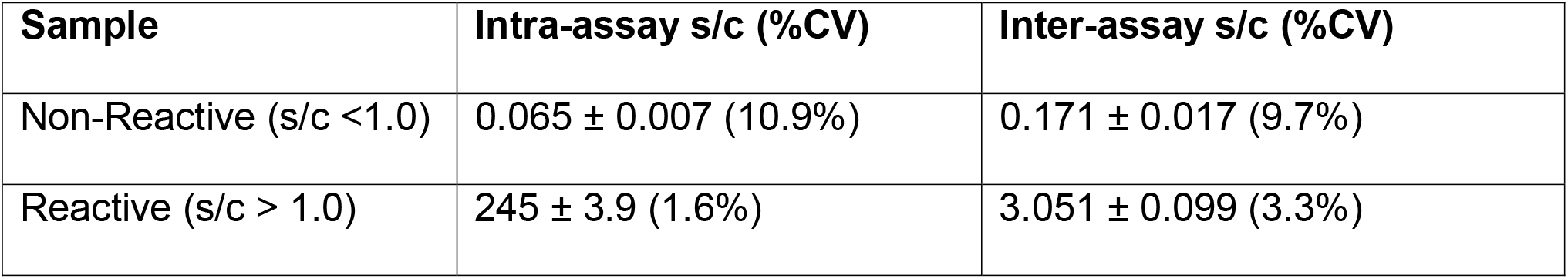
Intra- and inter-assay precision study results. S/c is given as mean ± standard deviation.

The CoV2T assay was concordant with negative and known positive cases of COVID-19 at our and neighboring institutions. Of healthy volunteer specimens, 100% (57/57) tested nonreactive by the CoV2T assay. Of SARS-CoV-2 PCR confirmed positive patient specimens, 70.9% (56/79) of all specimens tested reactive by CoV2T assay (Table 2). However, seroconversion in our patient population appeared to occur on day 4 to 7 after the onset of initial symptoms, and 96% of specimens collected on or after day 4 yielded a reactive result (Table 3). Only 6 of 17 (35%) specimens collected in the first three days of symptom onset were reactive by the CoV2T assay. Sensitivity (Positive predictive agreement or PPA) of the assay 7 days after of onset of symptoms was 100%, with the exception of one specimen which was reported to be PCR-positive at an outside location but was tested as negative in our hospital.

**Table 2.**
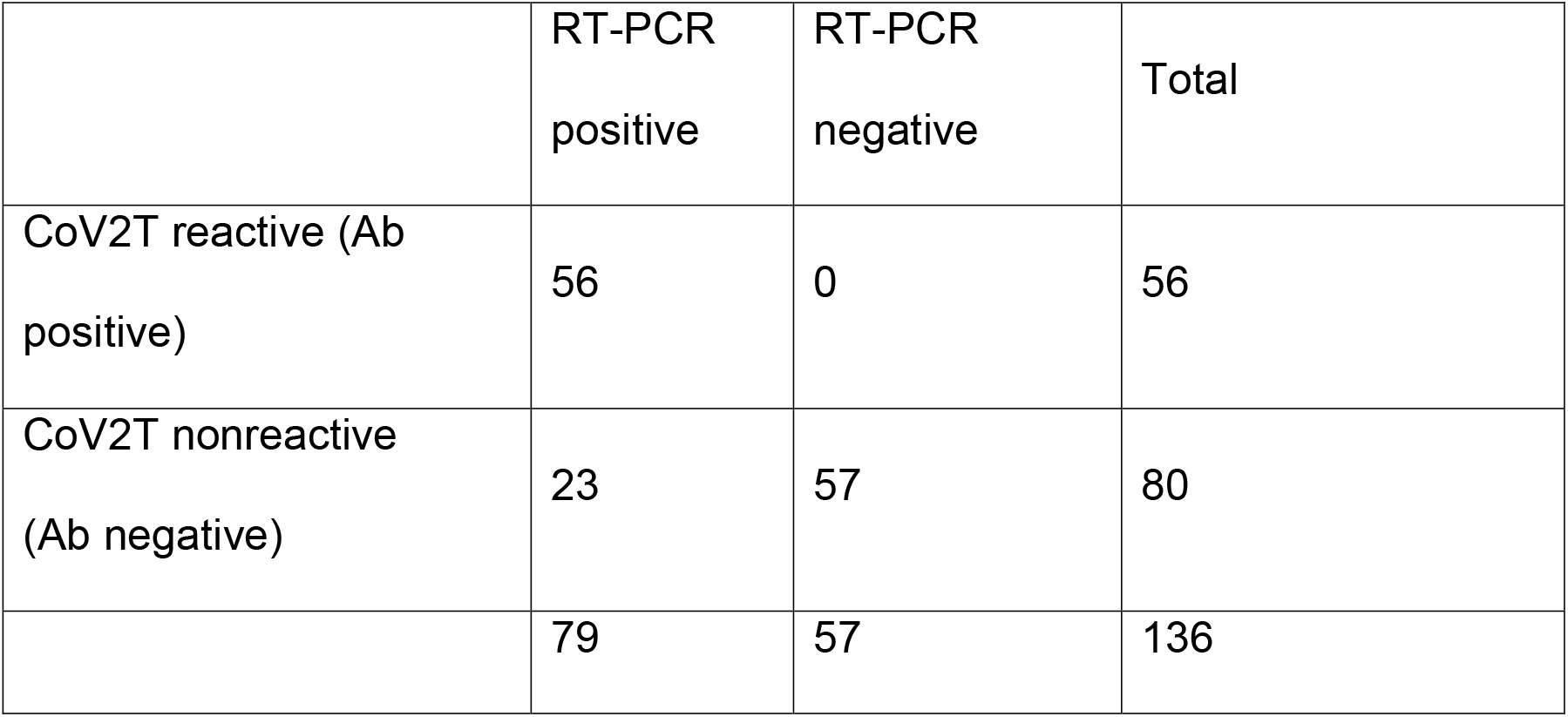
Concordance of the CoV2T assay with positive or negative results by RT-PCR for SARS-CoV-2.

|  | RT-PCR<br>positive | RT-PCR<br>negative | Total |
| --- | --- | --- | --- |
| CoV2T reactive (Ab<br>positive) | 56 | 0 | 56 |
| CoV2T nonreactive<br>(Ab negative) | 23 | 57 | 80 |
|  | 79 | 57 | 136 |

**Table 3.** Antibody reactivity by CoV2T assay in patients positive for SARS-CoV-2 by RT-PCR, grouped by number of days since the first reported symptom. Positive predictive agreement at > 7 days post-symptoms was 98%. *One patient in the >13 day group was reported positive at an outside hospital, but tested negative by RT-PCR at our institution.

| Days post-onset | N | Reactive |
| --- | --- | --- |
| < 3 | 17 | 35% (6/17) |
| 4-7 | 7 | 86% (6/7) |
| 8-13 | 8 | 100% (8/8) |
| > 13 | 23 | 96% (22/23)* |

There is no reference standard for serological testing of SARS-CoV-2, but we sought to compare the CoV2T assay with other serological methods for concordance. Twelve split specimens (4 nonreactive, 8 reactive) were tested at a reference laboratory using a qualitative assay, and were 100% concordant with the CoV2T assay. For the semiquantitative IgG method, the CoV2T assay was 91.2% concordant for negative samples (N = 80) and 91.6% concordant for positive samples (N = 48). (Table 4).

**Table 4.**
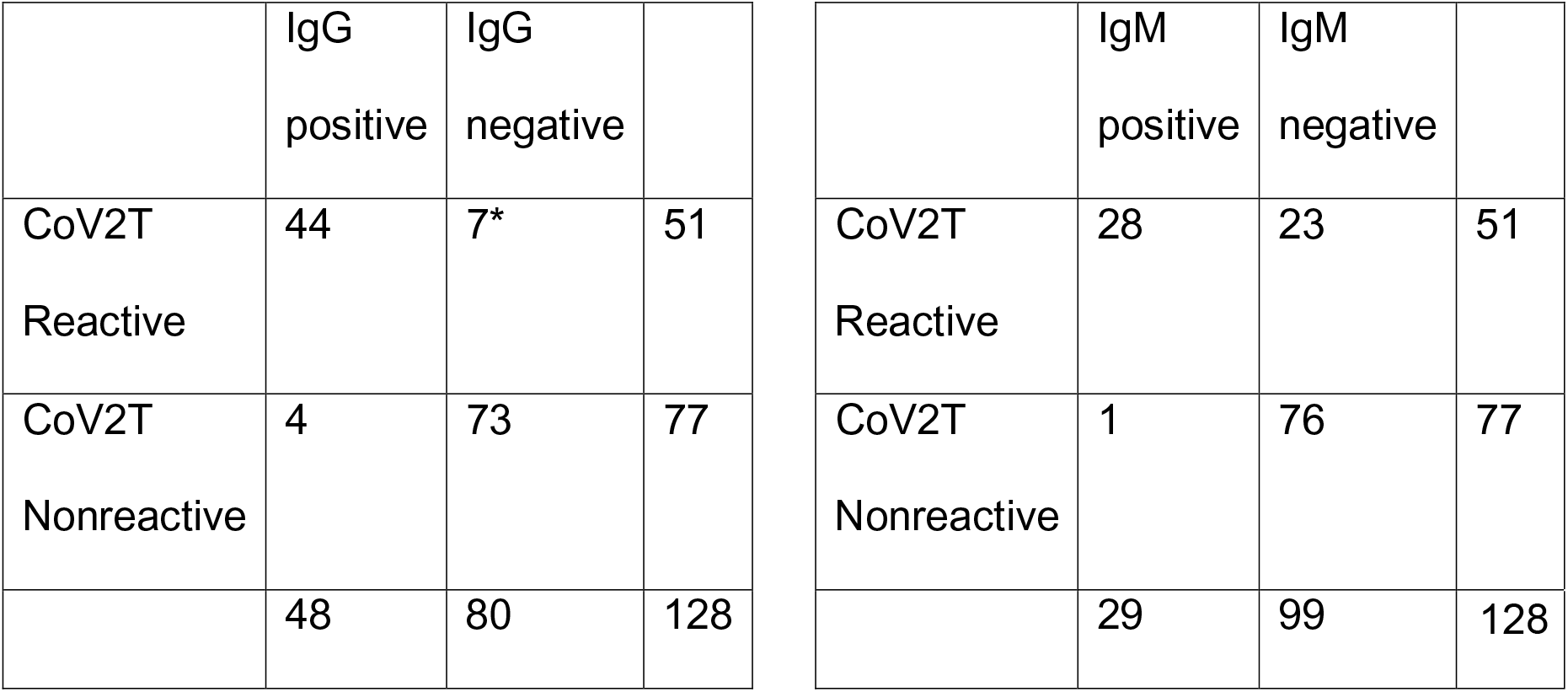
Method comparison with semiquantitative SARS-CoV-2 IgG and IgM ELISA. *Reactive specimens by CoV2T that were not IgG positive were not found to be positive for IgM by semiquantitative ELISA. All data from split sample testing with Viracor reference laboratories were concordant.

Analysis of plasma from patients with other viral infections did not indicate cross-reactivity with the CoV2T assay. Specimens from 14 patients previously tested to be negative for SARS-CoV-2 by RT-PCR, but positive for another respiratory viral infection by molecular analysis were nonreactive by the CoV2T assay. Interference studies identified no changes in sample reactivity when spiked with hemoglobin, conjugated bilirubin, or triglyceride-rich lipid (Table 5). Antibodies were not detected in specimens that were SARS-CoV-2 PCR negative (n=35), thereby yielding specificity (Negative Predictive agreement) of 100%.

**Table 5.**
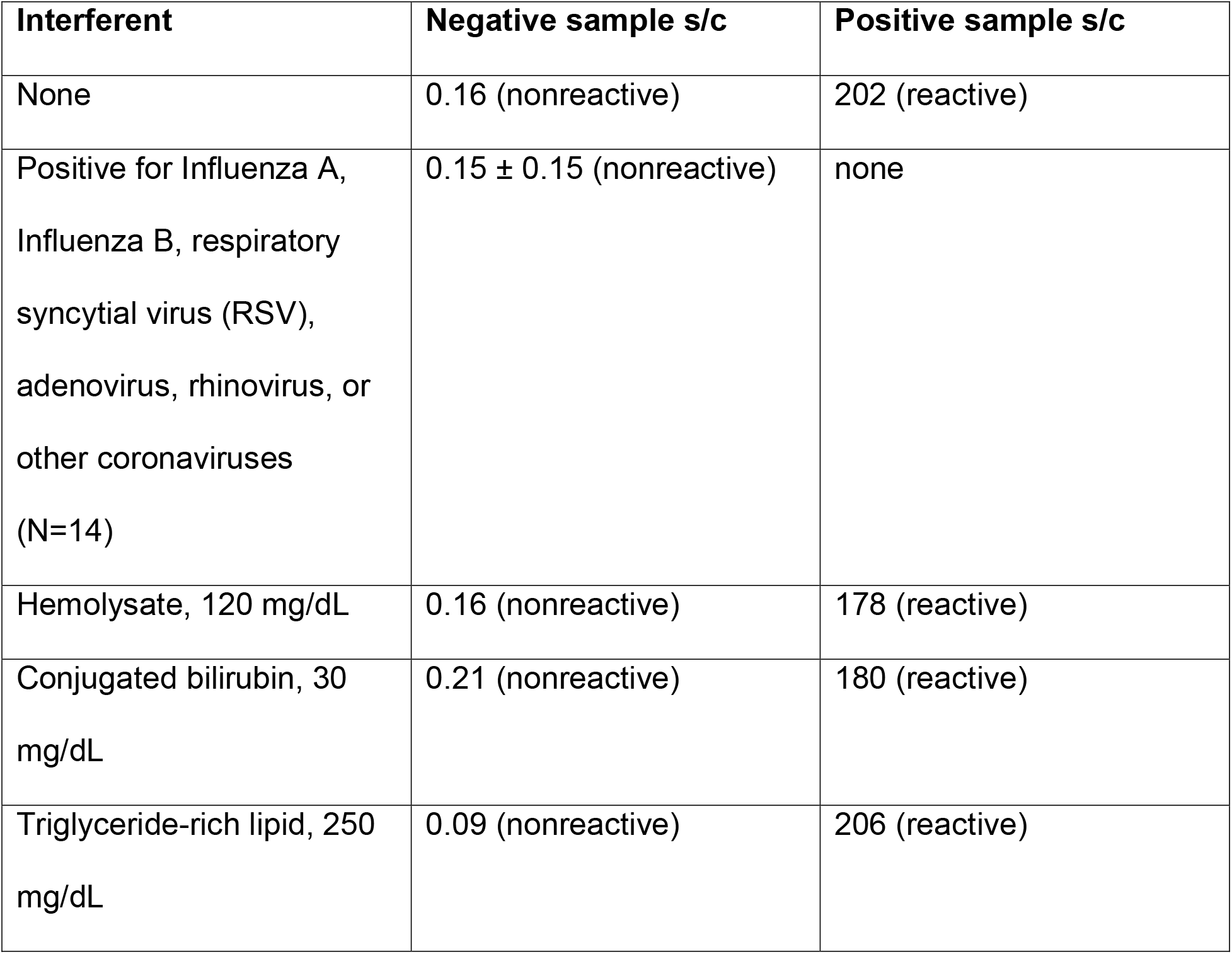
Analytical specificity studies with interference and other respiratory virus-positive specimens. For interference studies, known concentrations of interferent were spiked into either a known nonreactive or reactive sample. For other virus-positive specimens, patient specimens previously tested positive for another virus by molecular methods were tested by the CoV2T assay for reactivity.

The CoV2T instructions for use indicate that either serum or K_2_EDTA plasma may be used for analysis. Across five specimens, we verified that all specimens collected were concordant regardless of tube type used.

## Discussion

The availability of reliable serological testing for SARS-CoV-2 will be an important public health tool to mitigate the impact of COVID-19. Several serological assays have received an EUA by the FDA, and we describe here the validation of the Vitros CoV2T total antibody assay in our hospital laboratory. The CoV2T assay exhibited good analytical precision, good sensitivity (concordance with RT-PCR testing for SARS-CoV-2), and good concordance with other serological assays. We also confirmed that the assay had good specificity, and was robust to overcome common analytical interferences, and exhibited similar performance between serum and K_2_EDTA plasma samples. Our studies did not identify cross-reactivity of the CoV2T assay with other respiratory viruses or coronaviruses, but these studies were limited by the small number of available validation specimens in our laboratory. Further testing, particularly with banked specimens from prior to the emergence of SARS-CoV-2, will be useful to identify the possibility of cross-reactivity from other related viruses in this assay. Other studies have indicated low cross-reactivity of their serological assays with other coronaviruses using specimens from prior to the emergence of SARS-CoV-2, but this might differ slightly between assay manufacturers ^8^. Until the possibility of cross-reactivity is further evaluated, a positive antibody test by the CoV2T assay should be considered only presumptively positive for COVID-19 exposure.

Our studies also demonstrated excellent specificity (100%) with sensitivity of 70.9% in our complete validation sample set. The single specimen in our validation set that was discordant in a positive patient (positive by PCR at > 13 days post-onset, negative by CoV2T) was considered positive on the basis of an outside test result. When tested using our in-house RT-PCR assay, with a detection limit of as few as 40 copies of viral RNA per reaction, this specimen was negative for SARS-CoV-2. However, these data are preliminary, there is no standard reference method for serological testing to which to compare the CoV2T results, and the predictive value of the CoV2T assay when used as a screening test will be dictated by the actual prevalence of COVID-19 in our population and timing of specimen collection relative to the onset of symptoms. The performance of the CoV2T assay appears to be similar to that previously reported for other serological assays for COVID-19, with other methods internationally yielding sensitivity of 73.3-100% and specificity of 90.6-100% ^8-13^, however, performance characteristics are likely to vary based on epitope specificity and selected cutoff value for positive/reactive specimens. Further clinical performance studies are ongoing in our laboratory.

The higher specificity of the CoV2T assay relative to its sensitivity in our studies likely reflects the fact that a patient positive for SARS-CoV-2 by RT-PCR may be in the early stages of infection and may not yet be seropositive, resulting in false negatives. Studies of our patient population suggest that seropositivity as measured by the CoV2T assay may occur within the first 7 days, and show 97% sensitivity for SARS-CoV-2 at or beyond 7 days after the onset of symptoms. These data align with existing literature, which report that seropositivity for anti-SARS-CoV-2 antibodies occurs in only 50% of patients by 7 days after peak viral replication ^6,9^. The clinical utility of serological testing in management of COVID-19 patients is not yet known, but we concur that serological testing is likely to be of limited use for initial detection and diagnosis of symptomatic infections, largely due to variability in time between infection and detectable immune response. Additionally, in immunocompromised patients, any serological testing for SARS-CoV-2 assays may not be reliable. Consequently, molecular testing methods for viral RNA detection should be preferred as a diagnostic method.

Further, the CoV2T assay is qualitative, reporting total reactivity of IgG and IgM together rather than individually. Consequently, it cannot distinguish between convalescence and the acute phase of illness, two states that may be mistaken for one another given that many cases of COVID-19 have been reported to be asymptomatic. Use of an assay that measures IgG and IgM separately may be useful to identify convalescence, and confirmatory testing for by RT-PCR for SARS-CoV-2 would be necessary to rule out acute infection. Moreover, quantitative serological assays may be useful to evaluate associations between antibody titer and clinical classification.

Current FDA and CDC guidelines specify that while serological testing should not be used as a sole test to diagnose or rule out COVID-19, it may be useful to identify individuals with a history of recent exposure to SARS-CoV-2 ^7^. This information is important from a public health perspective both to identify the actual number of COVID-19 cases within a population (including those who were asymptomatic), and to determine whether a population has sufficient numbers of presumptively immune individuals to confer herd immunity.

Identification of actual case numbers permits generation of important epidemiological data, such as case fatality rates and correlation of demographic data with morbidity and mortality. Further, identifying individuals who are seropositive and recovered may be useful in treatment and prevention efforts. While it is yet unknown whether seropositivity against SARS-CoV-2 confers protection against reinfection with the virus, existing data suggest that reinfection is unlikely ^14^. Consequently, serological testing will likely be a useful tool for screening of workers, particularly essential workers along the front lines. In these individuals, serological testing may identify those at lower risk of infection from further exposure to COVID-19 and to inform who may return to work. However, given the limited information as of yet available on the clinical performance of this and other serological assays, it will be important to be vigilant for false positives in antibody testing. As with all clinical testing, the results from serological testing must be interpreted with respect to available epidemiological evidence and clinical context.

The use of convalescent plasma therapy in the treatment of COVID-19 has been authorized by the FDA for compassionate use ^15^. Convalescent plasma is derived from recovered and seropositive individuals, and thus, serological testing will be important to identify individuals who are eligible to donate plasma for this purpose. Antibodies from convalescent plasma may also be useful in the development of vaccines for SARS-CoV-2. In the future, serological tests can potentially be used in the evaluation of vaccine efficacy.

Future testing strategies for serological evaluation of SARS-CoV-2 may be able to specifically identify neutralizing antibody titers against the virus, as opposed to nonspecific or non-neutralizing antibodies. There is little existing information on the clinical relevance of neutralizing antibodies, but higher titers have been suggested to be correlated with milder disease course ^16^. The concordance of the CoV2T assay with methods to identify neutralizing antibodies will be important to know if clinically relevant.

There is also increasing evidence from the literature that serology testing may become very important in illnesses that occur in asymptomatic patients in the post-acute stage of COVID-19 infection, including unexpected thrombotic strokes and other thromboembolism, “COVID toes” and other cutaneous syndromes, and, as recently described, multi-inflammatory syndrome in the pediatric population ^17-19^. In these groups of patients, confirmation of previous exposure to SARS-CoV-2 by demonstration of seropositivity will be important to clearly link these disease states to COVID-19, which may help inform clinical management of patients.

In summary, we validated the Vitros CoV2T assay as a qualitative screening method for seropositivity against SARS-CoV-2 in our laboratory. We anticipate this assay will be a useful method for determination of true COVID-19 infection rates in our population, and may be useful as a predictor of immune status against COVID-19. While serological testing should be evaluated and implemented judiciously, it should ultimately provide a useful tool for surveillance and containment efforts in the ongoing COVID-19 pandemic.

## Data Availability

All data are provided in the manuscript

## Acknowledgement

EG and JJ were supported by the Ching Nan Ou Fellowship Endowment. Some of the validation kits used in this study were provided by Ortho Clinical Diagnostics, but they maintained no involvement in study design or validation, and were not privy to any of the data or interpretation.

